# Evaluation of high-throughput SARS-CoV-2 serological assays in a longitudinal cohort of mild COVID-19 patients: sensitivity, specificity and association with virus neutralization test

**DOI:** 10.1101/2020.09.30.20194290

**Authors:** Antonin Bal, Bruno Pozzetto, Mary-Anne Trabaud, Vanessa Escuret, Muriel Rabilloud, Carole Langlois-Jacques, Adèle Paul, Nicolas Guibert, Constance D’Aubarede-Frieh, Amélie Massardier-Pilonchery, Nicole Fabien, David Goncalves, André Boibieux, Florence Morfin-Sherpa, Virginie Pitiot, François Gueyffier, Bruno Lina, Jean-Baptiste Fassier, Sophie Trouillet-Assant, COVID SER STUDY GROUP

**Affiliations:** Laboratoire de Virologie, Institut des Agents Infectieux, Laboratoire associé au Centre National de Référence des virus des infections respiratoires, Hospices Civils de Lyon, Lyon, France; CIRI, Centre International de Recherche en Infectiologie, Team VirPath, Univ Lyon, Inserm, U1111, Université Claude Bernard Lyon 1, CNRS, UMR5308, ENS de Lyon, F-69007, Lyon, France; GIMAP EA 3064 (Groupe Immunité des Muqueuses et Agents Pathogènes), Université Jean Monnet, Lyon University, Saint-Etienne, France; Laboratory of Infectious Agents and Hygiene, University Hospital of Saint-Etienne, Saint-Etienne, France; Université de Lyon, F-69000, Lyon, France; Université Lyon 1, F-69100, Villeurbanne, France; Hospices Civils de Lyon, Pôle Santé Publique, Service de Biostatistique et Bioinformatique, F-69003, Lyon, France; CNRS, UMR 5558, University of Lyon, Laboratoire de Biométrie et Biologie Evolutive, Equipe Biostatistique-Santé, 69100, Villeurbanne, France; Lyon University, Université Claude Bernard Lyon1, Ifsttar, UMRESTTE, UMR T_9405, 8 avenue Rockefeller Lyon, France; Occupational Health and Medicine Department, Hospices Civils de Lyon, Lyon, France; Immunology Department, Lyon-Sud Hospital, Hospices Civils de Lyon, Pierre-Bénite, France; Infectious Diseases Department, Hospices Civils de Lyon, Lyon, France; Pharmacotoxicology Department, Hospices Civils de Lyon, Lyon, France

**Keywords:** COVID-19, SARS-CoV-2, Serological assays, Virus neutralization, Health-care workers

## Abstract

**Background:** The association between SARS-CoV-2 commercial serological assays and virus neutralization test (VNT) has been poorly explored in mild COVID-19 patients.

**Methods:** A total of 439 serum specimens were longitudinally collected from 76 healthcare workers with RT-PCR-confirmed COVID-19. The sensitivity (determined weekly) of nine commercial serological assays were evaluated. Specificity was assessed using 69 pre-pandemic sera. Correlation, agreement and concordance with the VNT were also assessed on a subset of 170 samples. Area under the ROC curve (AUC) was estimated at several neutralizing antibody titers.

**Results:** The Wantai Total Ab assay targeting the receptor binding domain (RBD) within the S protein presented the best sensitivity at different times during the course of disease. The specificity was greater than 95% for all tests except for the Euroimmun IgA assay. The overall agreement with the presence of neutralizing antibodies ranged from 62.2% (95%CI; 56.0-68.1) for bioMérieux IgM to 91.2% (87.0-94.2) for Siemens. The lowest negative percent agreement (NPA) was found with the Wantai Total Ab assay (NPA 33% (21.1-48.3)). The NPA for other total Ab or IgG assays targeting the S or the RBD was 80.7% (66.7-89.7), 90.3 (78.1-96.1) and 96.8% (86.8-99.3) for Siemens, bioMérieux IgG and DiaSorin, respectively. None of commercial assays have sufficient performance to detect a neutralizing titer of 80 (AUC<0.76).

**Conclusions:** Although some assays presented a better agreement with VNT than others, the present findings emphasize that commercialized serological tests including those targeting the RBD cannot substitute a VNT for the assessment of functional antibody response.

## Introduction

The evaluation of the humoral immune response to SARS-CoV-2 with serological tests is crucial to further manage the coronavirus disease 2019 (COVID-19) pandemic. Serological testing represents an easy to implement and cost-effective method allowing to rapidly identify individuals exposed to the virus (1,2). Over the last few months, a large number of SARS-CoV-2 commercial assays have been evaluated for their ability to detect specific antibodies (3–9). However, the detection of specific SARS-CoV-2 antibodies does not indicate whether or not the antibodies are functional for neutralizing the virus. In association with the assessment of other immune responses, such as cellular immunity, the exploration of the neutralizing antibody response is important to evaluate the protective immunity to SARS-CoV-2 after infection and therefore the risk of reinfection (10–13). To date, the association between SARS-CoV-2 commercial assay results and the presence of neutralizing antibodies has been mainly explored in hospitalized COVID-19 patients (14–18). As conflicting findings were reported, it is unclear whether the commercial serological assays would be useful to assess the protective immunity against SARS-CoV-2.

Virus neutralization test (VNT) is considered as the reference to assess the functional ability of antibodies to block the entry of the virus into human cells (19). However, such an assay requires living virus manipulated in a biosafety level 3 (BSL3) facility that needs trained staff and specific equipment, and which is a tedious and time-consuming method. The first study exploring the association of commercial serological assays and VNT claimed that the Wantai Total Ab assay detecting total antibodies directed against the SARS-CoV-2 receptor binding domain (RBD) had the best characteristics to detect functional antibodies at different stages and severity of disease (14). The RBD, within the sub-unit S1 of the spike protein, enables the viral entry into human cells by fixing to the angiotensin-converting enzyme 2 (ACE2) receptor (20). As emphasized by the authors (14), there is an urgent need for further studies addressing the performance of alternative high-throughput assays in correlation with VNT among persons with mild COVID-19 which is the most common form of the disease.

Thus, the aim of the present study was to evaluate widely-used SARS-CoV-2 serological tests and their potential association with VNT in a cohort of mild COVID-19 patients.

## Methods

### Study design and sample collection

A prospective longitudinal cohort study was conducted at the laboratory associated with the National reference center for respiratory viruses (University Hospital of Lyon, France)(21). Healthcare workers (HCW) with symptoms suggesting a SARS-CoV-2 infection requiring a RT-PCR test were included (visit 1, V1). Clinical data including date of symptom onset were recorded for all included HCWs using an eCRF by trained clinical research associate. Patients with a positive RT-PCR result at inclusion (V1) returned weekly for 6 additional visits (V2 toV7). Serum samples were collected at each visit. Written informed consent was obtained from all participants; ethics approval was obtained from the national review board for biomedical research in April 2020 (*Comité de Protection des Personnes Sud Méditerranée I*, Marseille, France; ID RCB 2020-A00932-37), and the study was registered on ClinicalTrials.gov (NCT04341142). A total of 439 serum specimens were longitudinally collected from 76 HCW. Among them, 74 had mild COVID-19 related symptoms (fever, cough, loss of taste or smell, diarrhea etc) and did not require hospital admission. Two out of 76 HCW were admitted to the hospital (not in intensive care unit, ICU) due to the severity of their symptoms. Among the 439 collected samples, 170 of them taken at V2, V4, V7 from 57 patients were tested by VNT (for one patient the sample at V7 was missing). To compare neutralizing antibody titers between mild and severe COVID-19 patients, 117 sera collected longitudinally from 44 severe COVID-19 patients admitted to ICU were also tested by VNT. In addition, to evaluate specificity we selected retrospectively 69 sera collected (before August 2019) from 30 healthy volunteers, 30 patients with autoimmune diseases, and 9 patients with a positive serological results for *M. pneumonia*e.

### Virological investigation

COVID-19 diagnosis at inclusion was performed by RT-PCR on nasopharyngeal swab using the cobas SARS-CoV-2 assay (Roche, Basel, Switzerland).

A total of 9 serological assays (Abbott, DiaSorin, Siemens, Bio-Rad, Wantai Total and IgM, bioMérieux IgG and IgM, Euroimmun IgA) were investigated according to the protocol recommended by each manufacturer (characteristics are summarized in Table 1). Positivity was established according to threshold value recommended by each manufacturer.

**Table 1.** Sensitivity of 9 SARS-CoV-2 commercial serological assays. Positivity was established according to threshold value recommended by each manufacturer. Ab: antibodies, Ig: immunoglobulin, ELISA: enzyme-linked immunosorbent assay, CMIA: chemiluminescence microparticule immune assay CLIA: chemiluminescence immune assay, ELFA: enzyme-linked fluorescent assay, n: number of samples, CI: confidence interval.

| Manufacturer (platform) | Abbott (Architect) | DiaSorin (Liaison®) | Siemens (Atellica®) | Bio-Rad | Wantai |  | bioMérieux (Vidas®) |  | Euroimmun |
| --- | --- | --- | --- | --- | --- | --- | --- | --- | --- |
| Assay name | SARS-CoV-2 IgG | SARS-CoV-2 S1/S2 IgG | SARS-CoV-2 Total | Platelia SARS-CoV-2 Total Ab | SARS-CoV-2 Total Ab | SARS-CoV-2 IgM | SARS-CoV-2 IgG | SARS-CoV-2 IgM | SARS-CoV-2 IgA |
| Assay type | CMIA | CLIA | CLIA | ELISA | ELISA | ELISA | ELFA | ELFA | ELISA |
| Antigen | N | S1+S2 | RBD | N | RBD | RBD | RBD | RBD | S1 |

Table 1 – Sensitivity of 9 SARS-CoV-2 commercial serological assays.
|  |  |  |  |  |  |  |  |  |  |
| --- | --- | --- | --- | --- | --- | --- | --- | --- | --- |
| days after symptom onset (n) |  |  |  |  |  |  |  |  |  |
| [1-7] (61) | 9.84 [5.17-17.91] | 6.56 [2.98-13.83] | 6.56 [2.98-13.83] | 18.03 [11.35-27.43] | 22.95 [15.36-32.84] | 13.11 [7.55-21.81] | 8.20 [4.05-15.90] | 11.48 [6.34-19.88] | 25.00 [17.02-35.14] |
| [8-14] (63) | 59.68 [49.23-69.31] | 32.26 [23.41-42.59] | 41.94 [32.18-52.37] | 74.19 [64.18-82.19] | 79.03 [69.41-86.23] | 64.52 [54.11-73.71] | 39.68 [30.17-50.04] | 49.21 [39.09-59.38] | 72.58 [62.47-80.81] |
| [15-21] (59) | 91.53 [83.60-95.81] | 83.05 [73.61-89.59] | 89.83 [81.52-94.65] | 96.61 [90.26-98.87] | 100.00 [95.62-100.00] | 94.92 [87.94-97.95] | 86.44 [77.50-92.19] | 77.97 [67.97-85.50] | 96.61 [90.26-98.87] |
| [22-28] (59) | 93.22 [85.73-96.92] | 86.44 [77.50-92.19] | 93.22 [85.73-96.92] | 94.92 [87.94-97.95] | 100.00 [95.62-100.00] | 89.83 [81.52-94.65] | 93.22 [85.73-96.92] | 69.49 [58.96-78.32] | 91.53 [83.60-95.81] |
| [29-35] (65) | 86.15 [77.66-91.76] | 92.31 [85.03-96.21] | 93.85 [86.98-97.21] | 92.19 [84.81-96.15] | 100.00 [96.00-100.00] | 84.62 [75.89-90.58] | 90.77 [83.13-95.15] | 52.31 [42.23-62.20] | 84.62 [75.89-90.58] |
| [36-42] (59) | 89.83 [81.52-94.65] | 93.22 [85.73-96.92] | 98.31 [92.75-99.62] | 91.53 [83.60-95.81] | 100.00 [95.62-100.00] | 88.14 [79.49-93.44] | 94.92 [87.94-97.95] | 45.76 [35.51-56.38] | 88.14 [79.49-93.44] |
| [43-85] (73) | 89.04 [81.58-93.71] | 89.04 [81.58-93.71] | 95.89 [90.15-98.35] | 88.89 [81.34-93.62] | 100.00 [96.38-100.00] | 81.94 [73.38-88.20] | 87.67 [79.97-92.68] | 43.84 [34.67-53.44] | 79.45 [70.69-86.11] |

A plaque reduction neutralization test (PRNT) was used for the detection and titration of neutralizing antibodies, as previously described (22). Briefly, a ten-fold dilution of each serum specimen in culture medium (Dulbecco’s Modified Eagle Medium containing antibiotics and 2% foetal calf serum) was first heated for 30 min at 56°C to avoid complement-linked reduction of the viral activity. Serial two-fold dilutions (tested in duplicate) of the serum specimens in culture medium were mixed at equal volume with the live SARS-CoV2 virus. After gentle shaking and a contact of 30 minutes at room temperature in plastic microplates, 150 µL of the mix was transferred into 96-well microplates covered with Vero E6 cells. The plates were incubated at 37°C in a 5% CO_2_ atmosphere. The reading was evaluated microscopically 5 to 6 days later when the cytopathic effect of the virus control reached 100 TCID_50_/150 µL. Neutralization was recorded if more than 50% of the cells present in the well were preserved. The neutralizing titer was expressed as the inverse of the higher serum dilution that exhibited neutralizing activity; a threshold of 20 was used (PRNT_50_ titer ≥ 20). All experiments were performed in a BSL3 laboratory. The comparison of this VNT with a standardized assay using retroviruses pseudo-typed with the SARS-CoV-2 S viral surface protein found a high correlation and concordance (22).

### Statistical analyses

For each test the clinical sensitivity was estimated weekly after symptom onset considering SARS-CoV-2 RT-PCR results as the gold standard.

Regarding VNT, the overall, positive and negative percent agreements (OPA, PPA, NPA) were determined for each commercial serological assay as previously described (23). The correlation and concordance with the VNT were assessed using the Spearman and Cohen’s Kappa coefficients, respectively. The concordance was classified as slight (Cohen’s Kappa coefficient, [0-0.2]), fair [0.21-0.4], moderate [0.41-0.6], substantial [0.61-0.8], and almost perfect [0.81-1] according to Landis and Koch criteria (24). The Cohen’s Kappa coefficient was not interpreted if the sensitivity was 100%. The estimation of the correlation coefficient was not performed due to an upper limit of signal to cut-off ratio for the Siemens and Bio-Rad assays. Specificity was assessed with 69 pre-pandemic serum specimens collected in 2019. The estimates are given with their bilateral 95% confidence interval (CI) calculated using the Wilson method. The 95% CI for Cohen’s Kappa coefficient was calculated using the bootstrap percentile method. The paired comparison of sensitivity between two assays was performed with the non-parametric McNemar test. The area under the ROC curve (AUC) was estimated to assess the overall performance of serological assays to detect the presence of neutralizing antibodies (PRNT_50_ ≥ 20) or higher neutralizing antibody titers (PRNT_50_ ≥ 80). Statistical analyses were carried out using SAS software, version 9.4 (Copyright (c) 2002-2003 by SAS Institute Inc., Cary, NC, USA.) and R software, version 3.6.1 (R Foundation for Statistical Computing, Vienna, Austria). A p-value < 0.05 was considered as statistically significant.

## Results

### Sensitivity and specificity

During the first week after the onset of symptoms the sensitivity for the detection of SARS-CoV-2 antibodies ranged from 6.6% (DiaSorin, Liaison) to 25.0% (Euroimmun IgA). The second week the sensitivity was greater than 70% for three tests including Bio-Rad, Wantai Total Ab, and Euroimmun IgA assays (74.2%, 79.0% and 72.6%, respectively). The highest of sensitivity was found at week # 3 for Bio-Rad (96.6%), Wantai Total Ab (100%), Wantai IgM (94.9%), bioMérieux IgM (78.0%) and Euroimmun IgA (96.6%), at week # 4 for Abbott (93.2%), and at week # 6 for DiaSorin (93.2%), Siemens (98.3%) and bioMérieux IgG (94.9%). After this point, a decrease of sensitivity was noted for all assays except for the Wantai Total Ab which remained steady at 100% over the course of the disease (Table 1). The Wantai Total Ab assay had a significantly higher sensitivity before 14 days post-symptom onset with all other assays, except with the Euroimmun IgA and Bio-Rad assays; after 14 days post-symptom onset, the differences were significant with all other assays.

In addition, we evaluated the specificity using 69 pre-pandemic sera. No false positive result was found with the two Wantai assays, bioMérieux IgG, and Siemens assays. For the other assays tests, the specificity was higher than 95% except for the Euroimmun IgA assay (84.06%, 95%CI [72.84-91.40]). Supplementary Table 1).

### Kinetics of neutralizing antibody titers

The neutralizing capacity of antibodies was determined at three time points for 57 patients (n=170 samples, Figure 1). No neutralizing antibodies were detected in 42.0% (21/50), 5.8% (3/51), and 8.7% (6/69) of samples collected between, respectively, 1-14, 15-28, and more than 28 days after symptom onset. Of note, three out of 57 patients had no neutralizing antibodies throughout their follow-up.

**Figure 1.**
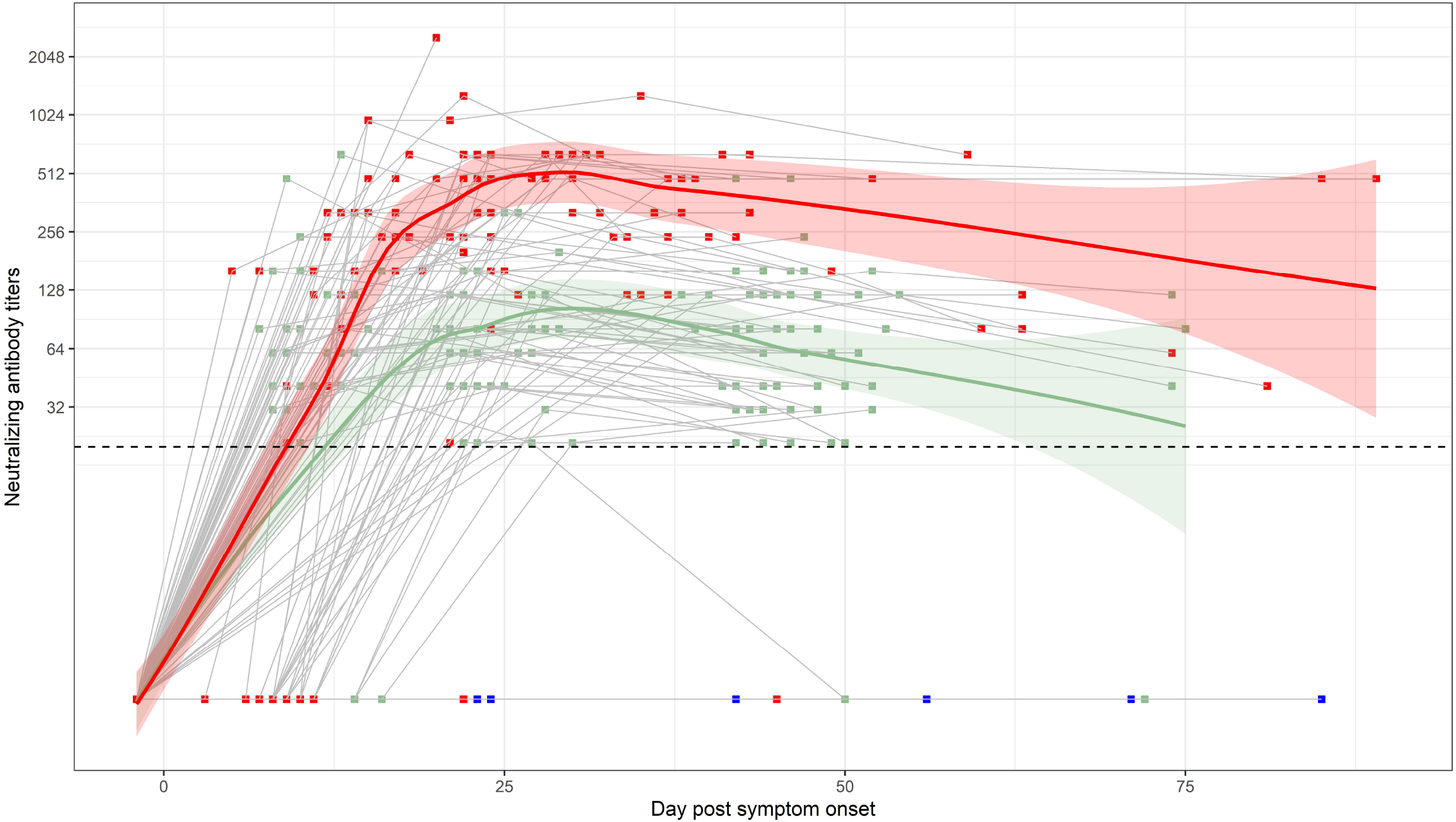
Kinetics of neutralizing antibody titers in mild and severe COVID-19 patients according to the post-symptom interval. Green points represent mild COVID-19 patients, red points represent severe COVID-19 patients admitted to ICU, and blue points represent patients without neutralizing antibodies throughout follow-up. Dotted lines correspond to the limit of quantification of neutralizing antibodies. Fit Loess curve represents local polynomial regression performed using the Loess method. CI at 95% is indicated (grey area).

For comparison, we also determined the titers of neutralizing antibodies in sera longitudinally collected from COVID-19 patients (n=44) admitted to an ICU (n=117 samples, Figure 1). Only one patient had no neutralizing antibodies throughout follow-up (until 45 days post symptoms). No neutralizing antibodies were detected in 42.3% (11/26), 2.3% (1/44), and 2.1% (1/47) of samples collected between, respectively, 1-14, 15-28, and more than 28 days after symptom onset.

For the samples with a detection of neutralizing antibody (n=140 and n=104 for mild and severe COVID-19 patients, respectively), the median [IQR] titer was 60 [40-100] vs 160 [80-320] between 1-14 days post symptom, reached 80 [60-120] vs 480 [240-640] between 15-28 days post symptom and decreased in samples collected after more than 28 days (median: 60 [40-120] vs 320 [120-640]).

### Comparison of results between commercial kits and VNT

The Spearman coefficient [95%CI] assessing correlation between commercial kits and VNT varied from 0.43 [0.27-0.56] to 0.61 [0.49-0.71] (Figure 2, Table 2).

**Table 2.** Association of SARS-CoV-2 commercial serological and a virus neutralization test. Ab: antibodies, Ig: immunoglobulin, ELISA: enzyme-linked immunosorbent assay, CMIA: chemiluminescence microparticule immune assay CLIA: chemiluminescence immune assay, ELFA: enzyme-linked fluorescent assay, n: number of samples, CI: confidence interval, dps: days post onset of symptoms, test. VNT: Virus neutralization test.

| Manufacturer (platform) | Abbott (Architect) | DiaSorin (Liaison®) | Siemens (Atellica®) | Bio-Rad | Wantai |  | bioMérieux (Vidas®) |  | Euroimmun |
| --- | --- | --- | --- | --- | --- | --- | --- | --- | --- |
| Assay name | SARS-CoV-2 IgG | SARS-CoV-2 S1/S2 IgG | SARS-CoV-2 Total | Platelia SARS-CoV-2 Total Ab | SARS-CoV-2 Total Ab | SARS-CoV-2 IgM | SARS-CoV-2 IgG | SARS-CoV-2 IgM | SARS-CoV-2 IgA |
| Assay type | CMIA | CLIA | CLIA | ELISA | ELISA | ELISA | ELFA | ELFA | ELISA |
| Antigen | N | S1+S2 | RBD | N | RBD | RBD | RBD | RBD | S1 |
| **Overall, Negative and Positive Percent Agreement with VNT** |  |  |  |  |  |  |  |  |  |
| OPA [95%CI] | 88.9 [84.3-92.3] | 89.5 [85.0-92.7] | 91.2 [87.0-94.2] | 89.4 [84.9-92.7] | 87.7 [82.9-91.2] | 81.2 [75.8-85.6] | 90.1 [85.7-93.3] | 62.2 [56.0-68.1] | 88.3 [83.7-91.8] |
| NPA [95%CI] | 74.2 [59.7-84.8] | 96.8 [86.8-99.3] | 80.7 [66.7-89.7] | 61.3 [46.6-74.2] | 33.3 [21.1-48.3] | 56.7 [41.9-70.4] | 90.3 [78.1-96.1] | 83.9 [70.4-91.9] | 67.7 [53.0-79.6] |
| PPA [95%CI] | 92.1 [87.6-95.1] | 87.9 [82.6-91.7] | 93.6 [89.3-96.2] | 95.7 [91.9-97.8] | 99.3 [96.9-99.9] | 86.4 [81.0-90.5] | 90.1 [85.1-93.5] | 57.4 [50.5-64.1] | 92.9 [88.4-95.7] |
| OPA [95%CI] <14dps | 85.4 [75.2-91.9] | 68.8 [57.0-78.5] | 83.3 [72.8-90.4] | 85.4 [75.2-91.9] | 75.0 [63.6-83.8] | 81.3 [70.4-88.8] | 69.4 [57.8-79.0] | 75.5 [64.3-84.1] | 81.3 [70.4-88.8] |
| OPA [95%CI] >14dps | 90.0 [84.6-93.7] | 97.5 [93.9-99.0] | 94.2 [89.6-96.8] | 91.6 [86.4-94.9] | 92.4 [87.4-95.6] | 83.2 [76.8-88.1] | 98.3 [95.1-99.5] | 58.3 [50.8-65.5] | 90.8 [85.6-94.3] |
| **Concordance with virus VNT - Cohen's Kappa coefficient [95%CI]** |  |  |  |  |  |  |  |  |  |
| Overall (n=170) | 0.64 [0.49-0.79] | 0.70 [0.56-0.83] | 0.72 [0.55-0.85] | 0.62 [0.44-0.76] | 0.43 [0.23-0.63] | 0.40 [0.21-0.58] | 0.71 [0.57-0.84] | 0.24 [0.14-0.36] | 0.61 [0.43-0.76] |
| <14 dps | 0.70 [0.45-0.88] | 0.41 [0.19-0.60] | 0.68 [0.45-0.84] | 0.69 [0.45-0.86] | 0.46 [0.21-0.67] | 0.61 [0.35-0.79] | 0.41 [0.29-0.76] | 0.52 [0.27-0.72] | 0.60 [0.35-0.79] |
| >14 dps | 0.40 [-0.06-0.72] | 0.86 [0.65-1] | 0.51 [0-0.89] | 0.45 [-0.05-0.79] | NA | 0.08 [-0.09-0.35] | 0.90 [0.71-1.00] | 0.18 [0.02-0.25] | 0.55 [0.06-0.81] |
| **Correlation between Ab level and neutralizing Ab titer** |  |  |  |  |  |  |  |  |  |
| Spearman coefficient [95%CI] | 0.47 [0.33-0.60] | 0.53 [0.39-0.65] | NA | NA | 0.56 [0.43-0.66] | 0.54 [0.40-0.65] | 0.61 [0.49-0.71] | 0.50 [0.32-0.65] | 0.43 [0.27-0.56] |
| **Receiver operating characteristic (ROC) curves for serology assays to detect neutralizing anti-SARS-CoV-2 antibodies or detect neutralizing - AUC [95%CI]** |  |  |  |  |  |  |  |  |  |
| PRNT50 ≥ 20 | 0.94 [0.89-0.98] | 0.96 [0.94-0.99] | 0.96 [0.94-0.99] | 0.85 [0.77-0.94] | 0.93 [0.88-0.98] | 0.80 [0.71-0.88] | 0.97 [0.94-0.99] | 0.80 [0.72-0.88] | 0.91 [0.85-0.97] |
| PRNT50 ≥ 80 | 0.65 [0.57-0.73] | 0.74 [0.66-0.81] | 0.73 [0.66-0.81] | 0.65 [0.58-0.72] | 0.75 [0.68-0.82] | 0.68 [0.60-0.76] | 0.75 [0.68-0.82] | 0.67 [0.58-0.75] | 0.64 [0.55-0.72] |

**Figure 2.**
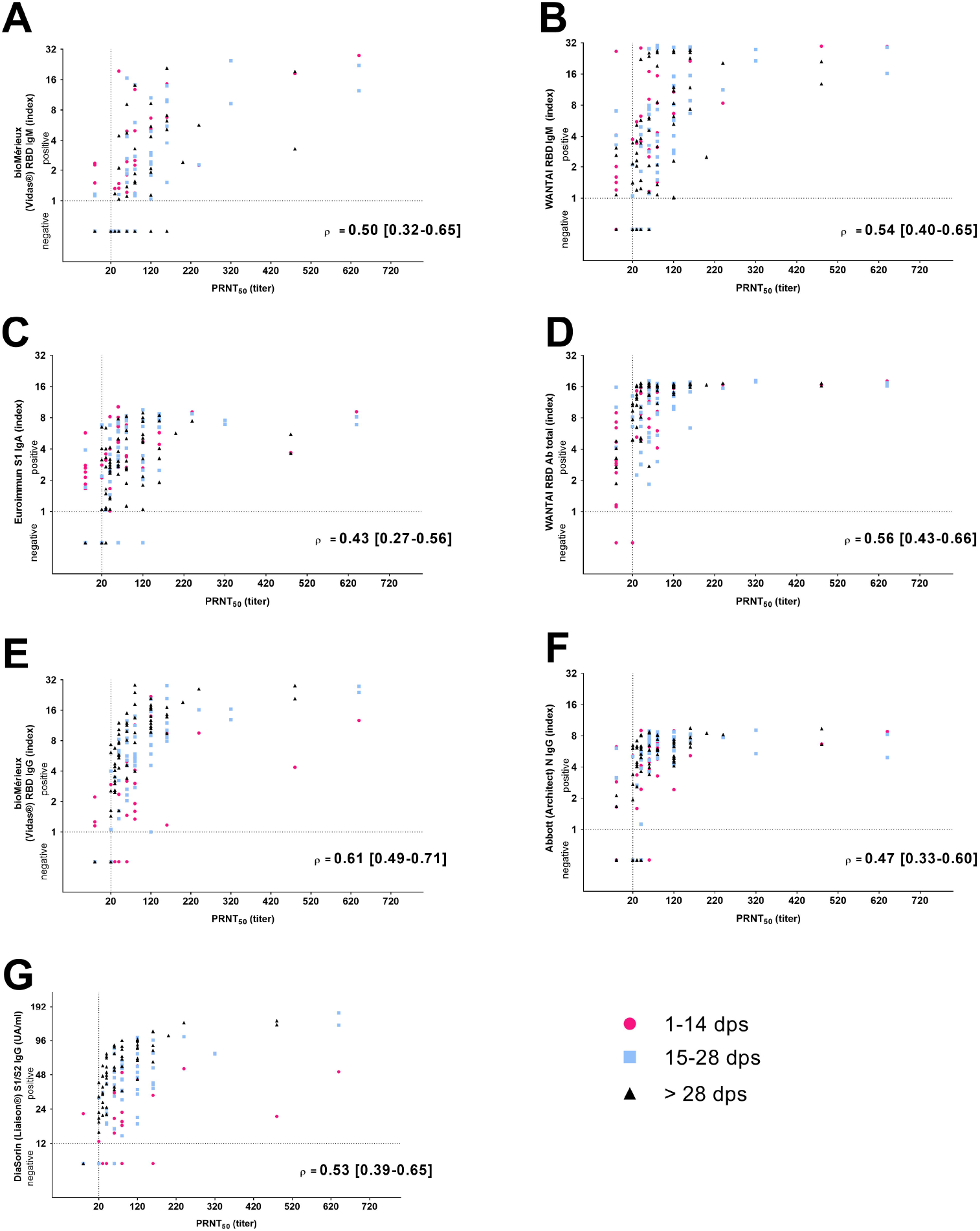
Correlation between SARS-CoV-2 neutralizing antibody titers and antibodies level determined by SARS-CoV-2 commercial serological assays. Magenta dots indicate sample collected ≤14 days post onset of symptoms (dps), blue dots indicate samples collected from 14-28 dps, black dots indicate specimen collected more than 28 dps. Spearman correlation coefficients and 95% confidence interval are indicated. All p-values were < 0.001.

A slight and fair concordance with VNT were noticed for the 2 IgM assays evaluated herein (Kappa [95%CI]: 0.24 [0.14-0.36] for bioMérieux IgM and 0.40 [0.21-0.58] for the Wantai IgM assays). Regarding total Ab or IgG assays targeting the S protein, three had substantial concordance with VNT (Kappa [95%CI]: 0.71[0.57-0.84] for bioMérieux, 0.70 [0.56-0.83] for DiaSorin, and 0.72 [0.55-0.85] for Siemens assays) while the concordance with the Wantai Total Ab assay was moderate (0.43 [0.23-0.63]; Table 2). The OPA, assessing the observed concordance between commercial serological assay and VNT confirmed that noticed with Cohen’s kappa. In particular, the lowest OPA was reported for the 2 IgM assays and for the Wantai Total Ab assay. Moreover, the NPA with VNT ranged from 33.3% [21.1-48.3] for the Wantai Total Ab assay to 96.8% [86.8-99.3] for the DiaSorin and was < 90% for 7/9 assays. The PPA with VNT was > 90% for all tests except the DiaSorin and the two IgM based assays (Wantai and bioMérieux) (Table 2).

Finally, ROC curves were built to estimate the performance of each commercial serological assay for detecting the presence of neutralizing antibodies (PRNT_50_ ≥ 20). The two IgM assays had the lowest AUC (0.80 for both). The AUC for the other assays found high performance to predict the presence of neutralizing antibodies reaching a value ≥ 0.96 for Siemens, Diasorin and bioMérieux IgG. The same methodology was applied for detecting higher neutralizing antibody titers (PRNT_50_ ≥ 80); none of these commercial assays had sufficient performance (AUC<0.76).

## Discussion

In a longitudinal study of 76 HCW with RT-PCR-confirmed COVID-19, we found that the Wantai Total Ab assay had the best sensitivity over the course of the disease. In particular, the sensitivity reached and remained at 100% as soon as week # 3 post symptom onset. This finding observed in mild COVID-19 patients is consistent with previous reports of excellent sensitivity of this test notably in severe patients (3,14). Importantly, the sensitivity of the commercial tests can be higher in severe patients in line with a stronger humoral immune response. In particular we found lower neutralizing antibody titers in mild patients than in ICU patients in the present study. These findings are highly consistent with previous studies (25–28) and raise questions about protective immunity after an infection although the immune response is not exclusively driven by the neutralizing antibody response. The immunological correlates of protection as well as the durability of natural immunity are still unknown, but mild-patients who had a low neutralizing antibody titer might be not enough protected to prevent a reinfection. The occurrence of reinfections in humans has been explored during a SARS-CoV-2 outbreak with a high attack rate which showed that individuals with preexisting neutralizing antibodies were not infected (10). Further studies are needed to investigate the correlation between neutralizing antibody titers and the risk of reinfections in mild COVID-19 patients. In addition, following the FDA recommendation regarding the required titer for convalescent plasma donors (titer ≥ 160), the data presented herein show that only few mild COVID-19 patients could be eligible. Thus, the ability of a commercial test to assess the neutralizing antibody response needs to be determined. With this aim, Tang et al. compared three commercial assays (Roche Total Ab, Abbott IgG, both tests targeting the N protein, and Euroimmun IgG assays targeting the S protein) to VNT on 66 specimens (15). They found that NPA was greater than 90% for all assays only at a low neutralizing titer of 20 while the NPA dramatically decreased when higher neutralizing titers were used, making them imperfect proxies for neutralization. For instance, the NPA for neutralizing titers was less than 60% for all 3 the assays at a cutoff of 128. Tang et al. also suggested to increase the manufacturer cutoff in order to improve the NPA which can be very useful in a vaccination setting or for plasma donor screening.

Although the study design was different, notably regarding the disease severity of the patients enrolled, these findings are highly consistent with those of the present study that found the lowest NPA for the Wantai Total Ab assay (33%) and a NPA below 90% for all tests except for bioMérieux IgG and DiaSorin. Importantly we also found with an AUC analysis that all the commercial tested performed poorly at a neutralizing antibody titer of 80.

Furthermore, the concordance between VNT and the Wantai Total Ab assay was only moderate while the concordance was substantial with bioMérieux IgG, DiaSorin, Siemens, Abbott, Euroimmun IgA and Bio-Rad. The low NPA and moderate concordance noticed for the Wantai Total Ab might be partially explained by the excellent ability of this test to detect RBD-specific antibodies at the very early phase of infection, irrespective of their neutralizing properties in line with the delay required for antibody maturation (29). In the first study comparing VNT with commercialized tests, the authors found that the Wantai Total Ab assay had the best characteristics to detect functional antibodies in different stages and severity of disease (14). However the median interval between the onset of symptoms and sample collection was 43 days for the mild patients samples tested (n=71 samples). Thus, the antibodies could be detected with both the Wantai Total Ab and the VNT assay at this time explaining the high PPA values noticed (14). Herein and in other reports (15–17), high PPA with VNT was also found for most of the commercial serological assays. Nevertheless, for determining the presence of neutralizing antibodies in serum specimen with commercial assays, the NPA should be maximized to avoid misinterpretation.

Furthermore, as previously reported by others (29–31), not all RBD-binding antibodies have neutralizing properties which is consistent with that reported herein regarding the RBD-based assays that do not have perfect concordance with VNT. Conversely, antibodies targeting a region other than the S protein may have functional activity, as previously reported (19,32– 34). In the present study, the Abbott and Bio-Rad assays directed against the N protein presented a substantial concordance with VNT. N-directed and RBD-neutralizing antibodies can be produced concomitantly over the course of the disease which can also explain this finding.

In addition to the different targeted antigens, the heterogeneity in assay performance found herein could be related to various factors including the detected isotypes. Moreover, antibody levels may also be very different according to the time since symptom onset and according to clinical severity of the disease (25). Herein, serum samples were collected longitudinally from disease diagnosis enabling to explore the early phase of the antibody response in a cohort of HCW, which constitutes one of the main strength of the present study.

The present study does, however, have certain limitations. For instance, specificity was not been extensively studied; yet the Euroimmun IgA assay seemed to have the worst specificity, which is consistent with previous studies reporting a lack of specificity for this assay (5,6,14). In addition, the performance of other notable commercial assays such as Euroimmun IgG or Roche Ig Total were not assessed. Second, not all the samples were systematically tested by VNT, in-line with the labor-intensive nature of this method. Finally, the size of the tested population remains small contributing to wide CI which limits the interpretation and extrapolation of the results.

The results presented herein confirm that, the Wantai Total Ab assay presented the higher sensitivity to detect SARS-CoV-2 antibodies after exposure. For the screening of neutralizing antibodies in serum specimens, optimized cut-off maximizing the NPA need to be established as previously suggest for the Wantai Total Ab assay (14). However, the data presented herein suggest that other tests targeting the S protein as Siemens, DiaSorin or bioMérieux IgG might be more useful for this indication.

These tests or others cannot substitute a VNT for assessing functional antibody response; neutralizing assays remain the gold standard and easy-to-use tests, such as those based on pseudoviruses (6,22,35), should be developed and standardized. Furthermore, the recent development of surrogate virus neutralization tests based on antibody-mediated blockage of the interaction between ACE-2 receptor and the RBD is very promising as they were designed in an ELISA format enabling high-throughput testing (30,36,37).

In conclusion, the present study provides original data concerning the performance of widely-used serological tests, which could help diagnostic laboratories in the choice of a particular assay according to the intended use.

### COVID-SER study group

Adnot Jérôme, Alfaiate Dulce, Bal Antonin, Bergeret Alain, Boibieux André, Bonnet Florent, Bourgeois Gaëlle, Brunel-Dalmas Florence, Caire Eurydice, Charbotel Barbara, Chiarello Pierre, Cotte Laurent, d’Aubarede Constance, Durupt François, Escuret Vanessa, Fascia Pascal, Fassier Jean-Baptiste, Fontaine Juliette, Gaillot-Durand Lucie, Gaymard Alexandre, Gillet Myriam, Godinot Matthieu, Gueyffier François, Guibert Nicolas, Josset Laurence, Lahousse Matthieu, Lina Bruno, Lozano Hélène, Makhloufi Djamila, Massardier-Pilonchéry Amélie, Milon Marie-Paule, Moll Frédéric, Morfin Florence, Narbey David, Nazare Julie-Anne, Oria Fatima, Paul Adèle, Perry Marielle, Pitiot Virginie, Prudent Mélanie, Rabilloud Muriel, Samperiz Audrey, Schlienger Isabelle, Simon Chantal, Trabaud Mary-Anne, Trouillet-Assant Sophie

## Data Availability

All data are available in the manuscript

## Acknowledgements

We thank all the personnel of the occupational health and medicine department of Hospices Civils de Lyon who contributed to the samples collection. We thank Lucie Charreton; Kahina Saker and Khadija Sfouli for their excellent work concerning serological testing, all the technicians from the virology laboratory whose work made it possible to obtain all these data, as well as Amira Lachekhab and Naima Rolnin for their technical assistance. We thank Karima Brahima and all members of the clinical research and innovation department for their reactivity (DRCI, Hospices Civils de Lyon). We thank Philip Robinson (DRCI, Hospices Civils de Lyon) for his help in manuscript preparation.

## Author contributor’s statement

All authors were involved in the analysis and interpretation of data as well as drafting the manuscript or revising it critically for important intellectual content. AB, BP, VP, FG, JBF and STA made substantial contributions to the conception and design of the study and designed the experiments. BP performed VNT. MAT and VE performed the serological assay experiments. NG, AP, CA, AMP, AB, and JBF were involved in patient care, VP performed the data collection, STA, AB, MA and BP performed the data analysis. MR and CLJ performed the statistical analysis. AB and STA wrote the paper, BL, BP, JBF, AP, VE and MAT revised the manuscript content. All authors read and approved the final manuscript.

## Conflict interests statement

Antonin Bal has received grant from bioMérieux and has served as consultant for bioMérieux for work and research not related to this manuscript. Sophie Trouillet-Assant has received research grant from bioMérieux concerning previous works not related to this manuscript. The other authors have no relevant affiliations or financial involvement with any organization or entity with a financial interest in or financial conflict with the subject matter or materials discussed in the manuscript.

## Funding

This research is being supported by Hospices Civils de Lyon and by Fondation des Hospices Civils de Lyon.

## Notes

### Competing Interest Statement

Antonin Bal has received grant from bioMerieux and has served as consultant for bioMerieux for work and research not related to this manuscript. Sophie Trouillet-Assant has received research grant from bioMerieux concerning previous works not related to this manuscript. The other authors have no relevant affiliations or financial involvement with any organization or entity with a financial interest in or financial conflict with the subject matter or materials discussed in the manuscript.

### Clinical Trial

NCT04341142

### Author Declarations

Written informed consent was obtained from all participants, approval was obtained from the national review board for biomedical research in April 2020 (Comite de Protection des Personnes Sud Mediterranee I, Marseille, France; ID RCB 2020-A00932-37)

